# Exploring Individual, Social, and Environmental Factors Related to Physical Activity: A Network Analysis

**DOI:** 10.1101/2024.03.22.24304703

**Authors:** Takeyuki Oba, Keisuke Takano, Kentaro Katahira, Kenta Kimura

## Abstract

**Objectives:** Insufficient physical activity (PA) has long been a global health issue, and a number of studies have explored correlates of PA to identify the mechanisms underlying inactive lifestyles. In the literature, dozens of correlates have been identified at different (e.g., individual, environmental) levels, but there is little or no direct evidence for the mutual associations of these correlates. This study analysed 44 variables identified as theoretically and empirically relevant for PA to clarify the factors directly and indirectly associated with PA.

**Methods:** A cross-sectional survey dataset of 19,005 Japanese-speaking adults (mean age = 53.50 years, SD = 17.40; 9,706 women) was analysed. The data encompassed demographic and anthropometric variables; self-reported PA levels; perceived social support and environments (e.g., awareness of urban facilities for PA); psychological traits and health-behaviour characteristics (e.g., personality, motivation, self-efficacy, decisional balance, process of change strategies); and technology use (e.g., mobile health apps).

**Results:** Network analyses were performed to select meaningful associations (partial correlations) among variables, which identified nine variables directly positively associated with PA: job/employment status, self-efficacy, perceived social support, intrinsic motivation, stage of change, counter conditioning, self-reevaluation, environment, and technology use. Indirect associations (two-step neighbourhood) were identified for 40 (out of 44) variables, implying that most of the kwon PA-correlates are associated with PA—at least indirectly.

**Conclusion:** Direct association with PA was identified for variables specified at different (individual–environmental) levels. The estimated mediation relationships echo the significance of the multilevel perspective in understanding how people maintain (in)active lifestyles.

**Summary Box:** *What is already known on this topic:*

- Research has identified numerous factors associated with physical activity (PA) to ascertain how people maintain physical inactivity and acquire healthier, more active lifestyles.
- Typically, empirical studies have investigated head-to-head associations between PA and PA-correlates, which do not, however, clarify the correlates’ mutual associations and the correlates that are directly and indirectly associated with PA.

*What this study adds:*

- We assessed 44 variables (across individual to environmental levels) that are known to be empirically and theoretically associated with PA among 19,005 Japanese-speaking adults,
- These variables were submitted to a psychological network analysis, which identified direct association with PA for the following variables (after controlling for all the other variables in the analysis): job/employment status, self-efficacy, perceived social support, intrinsic motivation for PA, stage of change, process-of-change strategies (counter conditioning and self-reevaluation), environment, and mobile-health-app use.
- Age and motivation for PA showed the highest centrality in the estimated network, implying that these two have the maximum number of associations with other correlates.

*How this study might affect research, practice, or policy:*

- The results highlight the significance of the multilevel approach—that is, understanding (in)active lifestyles from the perspective of individual characteristics (demographic, psychological, behavioural aspects, etc.) as well as social and environmental factors surrounding each individual.
- Our estimated network will guide stakeholders to specify the factors they should target in their projects (e.g., PA promotion) and estimate how those targets correlate with other factors that may lead to active lifestyles.

## Introduction

Insufficient physical activity (PA)—an unresolved issues in modern society ^1^—is a known risk factor for a variety of non-communicable and chronic diseases such as diabetes and cardiovascular and respiratory diseases ^2,3^. Researchers have endeavoured to identify the factors associated with PA from multiple perspectives. Demographic variables, such as age, gender, and health status, are robust predictors of PA levels ^4^. Psychological theories have highlighted the significance of motivation ^5^, self-efficacy ^6^, attitudes ^7^, and personality ^8^ in promoting PA. The rise of digital health, accelerated by the COVID-19 pandemic, has had a considerable impact on lifestyles as the use of smartphone apps and wearable activity trackers has been shown to be effective in increasing PA levels ^9^. Barriers and facilitators can also be identified among social, environmental, and political aspects surrounding individuals (e.g., support by family encouraging PA; access to walking trails; health programmes organised by local municipalities) ^10^.

Published review works have already provided a comprehensive overview of correlates and determinants of active lifestyles. Dishman et al.^11^ is one of the earliest, which extracted the factors contributing to regular PA from 41 research papers published during the 1970s and until the mid-1980s. They classified the extracted factors into the following three categories: personal (e.g., demographic and psychological factors), environmental (e.g., social support, peer influences), and activity characteristics (e.g., activity intensity). Trost et al.^12^ and Sallis and Owen ^13^ followed this line of research, reviewing empirical studies published during the 1990s. They expanded the taxonomy by adding a new category, physical environmental factors (e.g., adequate lighting, neighborhood safety), while updating the existing categories (e.g., dividing personal characteristics into demographic/biological, psychological, and behavioural factors). Bauman et al.^1^ echo the significance of the person-level (both psychological and biological) factors as well as social and physical environments, all of which can be located in a multilevel framework specifying PA correlates and determinants at different (individual, interpersonal, environment, policy, and global) levels. Although these literature reviews clarified the status of evidence and guided research on PA correlates, it remains vague how these factors are related with each other and which factors are directly and uniquely associated with PA. One of the most comprehensive lists of PA correlates is given by Trost et al. ^12^, which covers more than 70 factors extracted from published empirical studies that investigated the head-to-head associations with PA. Typically, those factors were studied separately in each empirical study – factor-to-factor associations are expected (e.g., individuals with agreeable personality may follow active peers encouraging PA, receive social support, and then acquire an active lifestyle) but technical challenges, particularly due to the number of identified factors, prevented researchers from drawing a full picture of the complex mediational associations around PA.

In the current study, we analysed 44 variables encompassing demographic, psychosocial, and environmental factors that are empirically and theoretically relevant for PA, using the psychological network analysis. This analytic approach enabled us to reveal the patterns of pairwise conditional dependencies present in a multivariate space (i.e., associations between the 45 variables: PA and 44 correlates) and to effectively visualise those patterns of statistical associations in the form of network diagram^14,15^. Our focus was on: (a) which factors would have a unique association with PA level (after controlling for the other factors in the data); (b) what mediational associations would emerge (or which factors would be indirectly associated with PA); and (c) which factors would be the most central in the network (having the greatest association with other variables in the network).

## Methods

### Participants

Participants (N = 20,611 Japanese speaking adults) were recruited from a sample-pool database; more than a million online panels had been registered to this database. The sample size was determined for practical and pragmatic reasons: (a) we expected that a large sample size would be required to estimate a network of 45 variables (and 990 possible associations), and therefore, we aimed for the maximum affordable number considering the financial and human resources that we had; and (b) the collected data were analysed for other purposes (not reported here), which specifically focused on a particular group of participants (e.g., mHealth app users) and required a sufficiently large size for the sub-sample. Eligible participants (i.e., being aged > 18 years, having a good command over Japanese, and residency in Japan) received an invitation for two online surveys separated by a month (early 2023). In the first survey, participants completed questionnaires regarding demographics, levels and readiness for PA, psychological characteristics, social support, and environmental factors relevant for PA. The second survey encompassed the current health status and medical histories. For each survey, participants received a small compensation for their participation (online shopping voucher). Of the 20,611 participants, 19,039 completed both the surveys. Data of 34 participants were deemed unreliable; that is, those reported: (1) a height of ≤ 100 cm; (2) a weight of ≤ 10 kg; and (3) total active time of > 24 hours per day. The remaining 19,005 responses were submitted for statistical analyses. All participants provided informed consent. This study was approved by the Ethics Committee of the National Institute of Advanced Industrial Science and Technology (Approval ID:2022-1279). We reported the results in accordance with Strengthening the Reporting of Observational Studies in Epidemiology (STROBE) statement ^16^.

### Equity, diversity and inclusion statement

Our team consisted of four Japanese men: one junior researcher and three senior researchers. The study population covered a wide range of age groups with balanced gender. However, the online survey was conducted in Japan exclusively and written in Japanese. We assumed that participants had good language command and internet literacy, which may have affected the demographics of participants and thus may limit the generalizability of the results (e.g., individuals with lower socioeconomic status or from more marginalised communities may not be included).

### Measures

Overall, we based variable selection on systematic literature reviews, such as Trost et al.^12^, which summarised and categorised PA-relevant factors into: demographic and biological; psychological, cognitive, and emotional; behavioural attributes and skills; social and cultural; physical environmental; and physical activity characteristics. Many of the variables listed here were assessed in the current study as well – Table 1 presents an overview of the demographic and anthropometric variables, PA levels and readiness, and health status (see also Table S1 for details regarding reported present diseases). Family incomes were binarised whether participant’s income was greater than the average family income in Japan (5 million yen). Education levels were also dichotomised to reflect whether participants graduated a university. Yet, there were several exceptions and deviations from Trost et al. ^12^. First, for psychological characteristics, we measured variables related to personality, motivation (regulatory focus; hedonic and eudaimonic motives for activities in general), self-control, and perceived social support (Table 2). We considered social support as a *psychological* characteristic as this was the only factor concerning social and cultural characteristics assessed in this study.

**Table 1:** Demographic and Descriptive Data (Total *N* = 19,005)

| Variable | Abbreviation <sup>a</sup> | Mean (SD), N (%) |
| --- | --- | --- |
| Age | Age | 53.50 (17.40) |
| Gender (women) | Gnd | 9706 (51.07%) |
| BMI | BMI | 22.15 (3.71) |
| Marital status (binary, yes/no) | Mrt | 12106 (63.70%) |
| Having a child (binary, yes/no) | Chl | 11860 (62.40%) |
| Education | Edc |  |
| Middle school |  | 462 (2.43%) |
| High school |  | 5876 (30.92%) |
| College or vocational school |  | 4302 (22.64%) |
| University or above |  | 8213 (43.21%) |
| Other |  | 152 (0.80%) |
| Job/Employment (binary, yes/no) | Emp | 11358 (59.76%) |
| Household income (JPY) | Inc |  |
| < 3 million |  | 4063 (21.38%) |
| 3 - 5 million |  | 4638 (24.40%) |
| 5 - 7 million |  | 2917 (15.35%) |
| 7 - 10 million |  | 2393 (12.59%) |
| 10 million or above |  | 1589 (8.36%) |
| No answer |  | 3405 (17.92%) |
| Total PA (in METs-hour/week) | PA | 35.06 (56.94) |
| Stage of change | SoC |  |
| Precontemplation |  | 4090 (21.5%) |
| Contemplation |  | 4168 (21.9%) |
| Preparation |  | 3687 (19.4%) |
| Action |  | 834 (4.39%) |
| Maintenance |  | 6226 (32.8%) |
| Health status |  |  |
| EQ5D-5L |  |  |
| Quality of life | QoL | 0.82 (0.15) |
| Health status (VAS, 0-100) | Hls | 76.32 (17.60) |
| Present disease (binary, yes/no) | Dss | 8605 (45.28%) |

Abbreviations: SD, Standard deviation; BMI, Body mass index; METs, Metabolic equivalents; VAS, visual analog scale
<sup>a</sup> Abbreviations used in the network diagram (Figures 1 and 2)

**Table 2:**
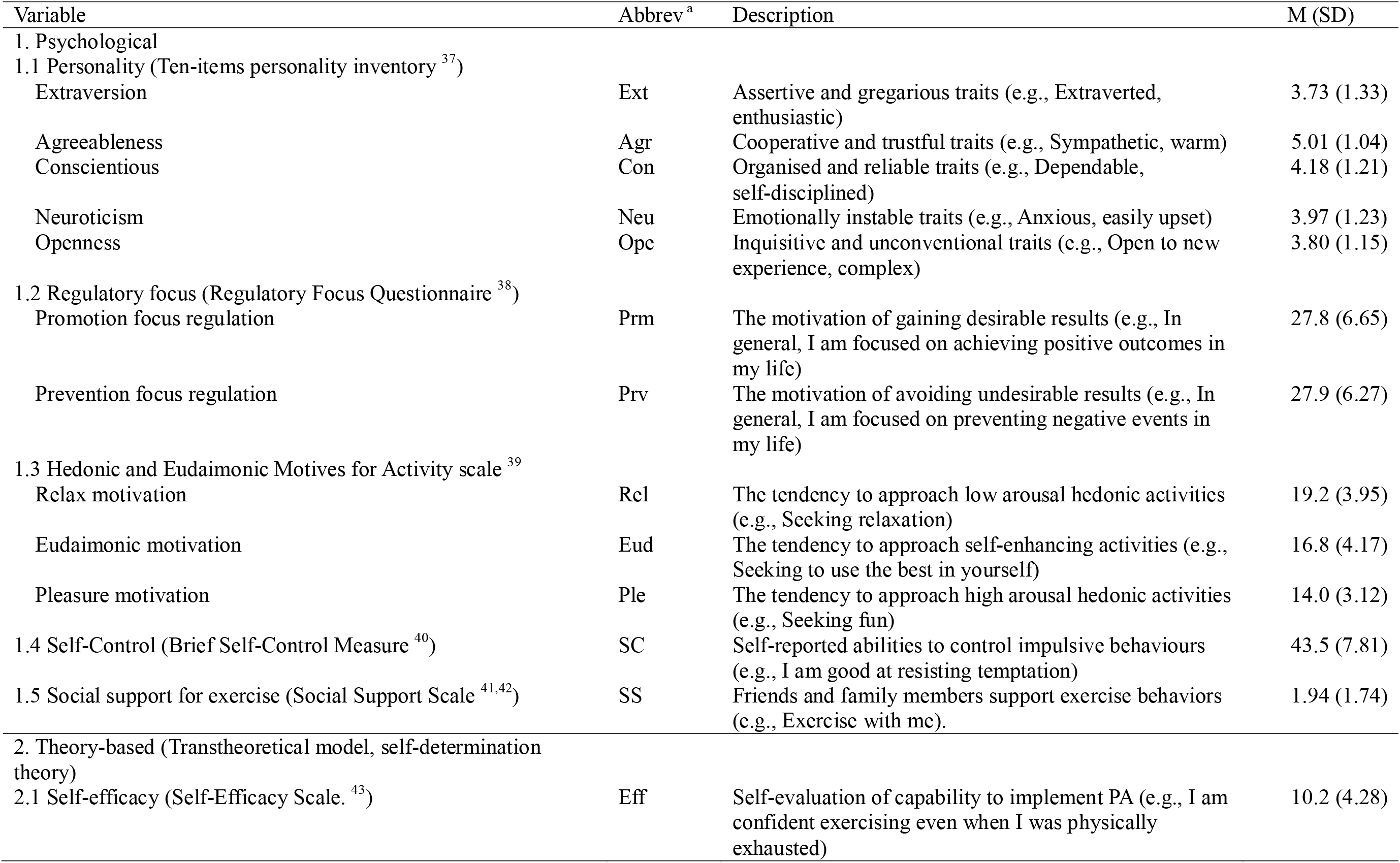
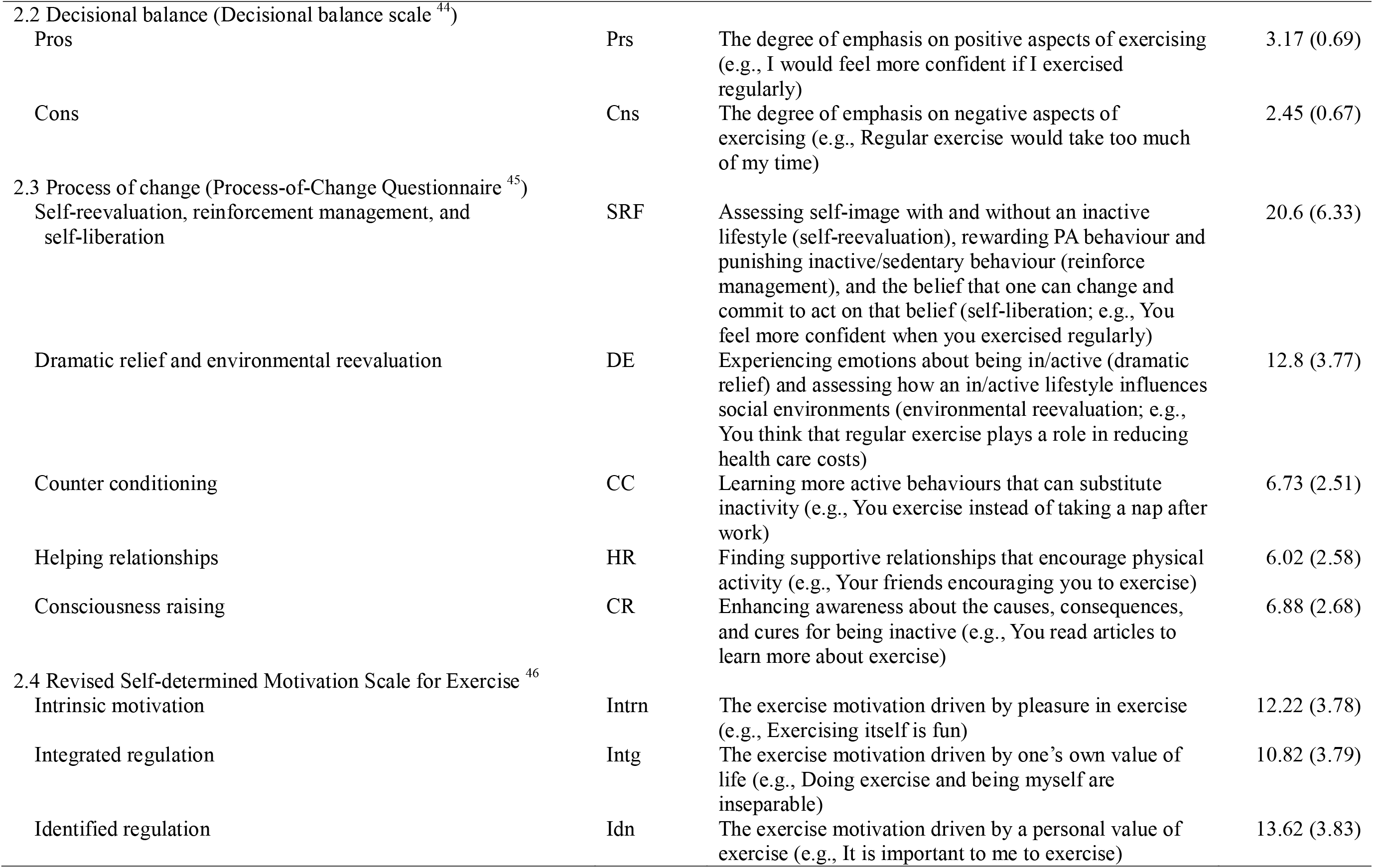
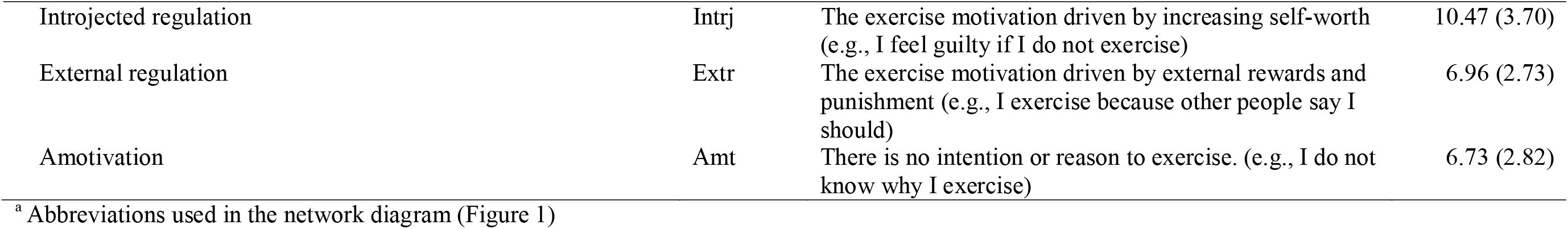
Psychological and Theory-based Variables.

| Variable | Abbrev<sup>a</sup> | Description | M (SD) |
| --- | --- | --- | --- |
| 1. Psychological |  |  |  |
| 1.1 Personality (Ten-items personality inventory <sup>37</sup>) |  |  |  |
| Extraversion | Ext | Assertive and gregarious traits (e.g., Extraverted, enthusiastic) | 3.73 (1.33) |
| Agreeableness | Agr | Cooperative and trustful traits (e.g., Sympathetic, warm) | 5.01 (1.04) |
| Conscientious | Con | Organised and reliable traits (e.g., Dependable, self-disciplined) | 4.18 (1.21) |
| Neuroticism | Neu | Emotionally instable traits (e.g., Anxious, easily upset) | 3.97 (1.23) |
| Openness | Ope | Inquisitive and unconventional traits (e.g., Open to new experience, complex) | 3.80 (1.15) |
| 1.2 Regulatory focus (Regulatory Focus Questionnaire <sup>38</sup>) |  |  |  |
| Promotion focus regulation | Prm | The motivation of gaining desirable results (e.g., In general, I am focused on achieving positive outcomes in my life) | 27.8 (6.65) |
| Prevention focus regulation | Prv | The motivation of avoiding undesirable results (e.g., In general, I am focused on preventing negative events in my life) | 27.9 (6.27) |
| 1.3 Hedonic and Eudaimonic Motives for Activity scale <sup>39</sup> |  |  |  |
| Relax motivation | Rel | The tendency to approach low arousal hedonic activities (e.g., Seeking relaxation) | 19.2 (3.95) |
| Eudaimonic motivation | Eud | The tendency to approach self-enhancing activities (e.g., Seeking to use the best in yourself) | 16.8 (4.17) |
| Pleasure motivation | Ple | The tendency to approach high arousal hedonic activities (e.g., Seeking fun) | 14.0 (3.12) |
| 1.4 Self-Control (Brief Self-Control Measure <sup>40</sup>) | SC | Self-reported abilities to control impulsive behaviours (e.g., I am good at resisting temptation) | 43.5 (7.81) |
| 1.5 Social support for exercise (Social Support Scale <sup>41,42</sup>) | SS | Friends and family members support exercise behaviors (e.g., Exercise with me). | 1.94 (1.74) |
| 2. Theory-based (Transtheoretical model, self-determination theory) |  |  |  |
| 2.1 Self-efficacy (Self-Efficacy Scale. <sup>43</sup>) | Eff | Self-evaluation of capability to implement PA (e.g., I am confident exercising even when I was physically exhausted) | 10.2 (4.28) |
| 2.2 Decisional balance (Decisional balance scale <sup>44</sup>) |  |  |  |
| Pros | Prs | The degree of emphasis on positive aspects of exercising (e.g., I would feel more confident if I exercised regularly) | 3.17 (0.69) |
| Cons | Cns | The degree of emphasis on negative aspects of exercising (e.g., Regular exercise would take too much of my time) | 2.45 (0.67) |
| 2.3 Process of change (Process-of-Change Questionnaire <sup>45</sup>) |  |  |  |
| Self-reevaluation, reinforcement management, and self-liberation | SRF | Assessing self-image with and without an inactive lifestyle (self-reevaluation), rewarding PA behaviour and punishing inactive/sedentary behaviour (reinforce management), and the belief that one can change and commit to act on that belief (self-liberation; e.g., You feel more confident when you exercised regularly) | 20.6 (6.33) |
| Dramatic relief and environmental reevaluation | DE | Experiencing emotions about being in/active (dramatic relief) and assessing how an in/active lifestyle influences social environments (environmental reevaluation; e.g., You think that regular exercise plays a role in reducing health care costs) | 12.8 (3.77) |
| Counter conditioning | CC | Learning more active behaviours that can substitute inactivity (e.g., You exercise instead of taking a nap after work) | 6.73 (2.51) |
| Helping relationships | HR | Finding supportive relationships that encourage physical activity (e.g., Your friends encouraging you to exercise) | 6.02 (2.58) |
| Consciousness raising | CR | Enhancing awareness about the causes, consequences, and cures for being inactive (e.g., You read articles to learn more about exercise) | 6.88 (2.68) |
| 2.4 Revised Self-determined Motivation Scale for Exercise <sup>46</sup> |  |  |  |
| Intrinsic motivation | Intrn | The exercise motivation driven by pleasure in exercise (e.g., Exercising itself is fun) | 12.22 (3.78) |
| Integrated regulation | Intg | The exercise motivation driven by one's own value of life (e.g., Doing exercise and being myself are inseparable) | 10.82 (3.79) |
| Identified regulation | Idn | The exercise motivation driven by a personal value of exercise (e.g., It is important to me to exercise) | 13.62 (3.83) |
| Introjected regulation | Intrj | The exercise motivation driven by increasing self-worth (e.g., I feel guilty if I do not exercise) | 10.47 (3.70) |
| External regulation | Extr | The exercise motivation driven by external rewards and punishment (e.g., I exercise because other people say I should) | 6.96 (2.73) |
| Amotivation | Amt | There is no intention or reason to exercise. (e.g., I do not know why I exercise) | 6.73 (2.82) |

<sup>a</sup> Abbreviations used in the network diagram (Figure 1)

Other psychological variables were grouped under the *theory-based* category together with behavioural variables (Table 2). These variables are typically drawn from the Transtheoretical model and Self-determination theory, encompassing motivation (specifically for exercise), self-efficacy, process of change, and decisional balance, which often tap into both psychological (cognitive) and behavioural aspects. As we wanted to distinguish between general psychological traits (e.g., personality, motivation in general) and the constructs specifically developed in the context of PA and exercise, we decided not to merge the general psychological and theory-based characteristics.

Like Trost et al. ^12^, we assessed physical environmental factors (Table 3), representing the presence of sidewalks and bike paths as well as safety from crime and traffic. We expected that access to *digital* (not only physical) resources would also be a key factor to explain PA, and thus, the use of mobile health technology (apps and wearable activity trackers supporting PA) and technology acceptance were added to the list (Table 3).

**Table 3:** Environmental and Technological Characteristics.

| Variable | Abbrev<sup>a</sup> | Description | Mean (SD),<br>N (%) |
| --- | --- | --- | --- |
| 1. Environment<br>(International Physical Activity Questionnaire Environmental Module) |  |  |  |
| Housing density | Hsd | Housing density in the neighbourhood (e.g., What is the main type of housing in your neighborhood? *Detached single-family residences*) | 2.06 (1.37) |
| Environment | Env | Evaluation that environment surrounding a house is suitable for exercise and walking (e.g., There is so much traffic on the streets that it makes it difficult or unpleasant to walk in my neighborhood) | 40.82 (6.28) |
| Number of vehicles | Veh | The number of vehicles (motorbikes or cars). | 1.33 (1.86) |
| 2. Technology |  |  |  |
| Use mHealth supporting PA | mHl | Currently using a smartphone app or wearable activity tracker supporting PA and exercise | 4587 (24.14%) |
| Technology acceptance<br>(Technology Acceptance Model scale <sup>47</sup>) |  |  |  |
| Usefulness of technology | Usf | Perception that technology tools help one's own works (e.g., Work more quickly) | 13.79 (3.48) |
| Ease of use of technology | Eas | Perception that technology tools are not difficult to use (e.g., Easy to become skillful) | 13.38 (3.42) |

<sup>a</sup> Abbreviations used in the network diagram (Figure 1)

### Statistical analysis

To find meaningful associations between the assessed variables, we estimated a network using the *qgraph* package ^17^ for R (version 4.2.2: R Core Team ^18^). This analysis estimated a Graphical Gaussian Model, in which an edge represents a partial correlation coefficient, indicating an association between a given pair of variables (nodes) after controlling for the other variables present in the network. Meaningful edges were selected by the graphical least absolute shrinkage and selection operator ^19^ (GLASSO). The best-fit network structure was searched using the Extended Bayesian Information Criteria ^20^ (EBIC) with the hyperparameter γ (a penalty term of the number of edges) set as 0.5 ^14^. Prior to network estimation, a non-paranormal transformation ^21^ was applied because the normality assumption was violated for certain (e.g., binary) variables (see the supplementary materials for a sensitivity analysis without the transformation, Figure S1). Each network diagram was depicted using the Fruchterman–Reingold algorithm^22^, which determines edge lengths depending on the absolute values of the edge weights (i.e., nodes with a higher partial correlation have a shorter edge length). On the estimated network, we first focused on the variables that had a direct association with PA. Second, we interpreted the indirect associations mediated by the direct nodes. Third, we computed two centrality indices: strength and expected influence, to explore how and which nodes were most closely associated with other nodes in the network. Strength centrality is given by the sum of absolute edge weights that a node has. Expected influence is the sum of signed weights, which considers the directions—positive and negative—of associations of a node ^23^. Edge accuracy (bootstrapped confidence intervals with 2.5 and 97.5% quantiles) and stability of the centrality indices ^24^ were assessed using the *<u>bootnet</u>* package (see the supplemental materials for the technical details, Figures S2 and S3).

## Results

Demographic information is presented in Table 1. The majority of participants were married (64%), had at least one child (62%), held a job (60%), and had obtained higher education (university or above; 43%). The mean BMI score was 22.15 (standard deviation = 3.71) and the total PA level was 35.06 METs-hours/week (standard deviation = 56.94). As 9,500 participants (50%) met the levels of PA recommended by the national guideline (i.e., 23 METs-hours/week for adults aged < 65 years; 10 METs-hours/week for older ^25^), our sample might be more active than the general population.

The estimated network is illustrated in Figure 1A, presenting all 295 edges selected by the GLASSO algorithm. Panel B specifically illustrates the edges directly associated with PA (one-step neighborhood). Of the 44 PA-correlates submitted to the network analyses, nine showed positive, direct associations: *job/employment*, edge weight = 0.049, IQR = [0.036, 0.062]; *stage of change*, edge weight = 0.256, IQR = [0.242, 0.270]; *social support*, edge weight = 0.073, IQR = [0.059, 0.087]; *self-efficacy*, edge weight = 0.062, IQR = [0.048, 0.077]; *self-reevaluation*, edge weight = 0.024, IQR = [0, 0.037]; *counter conditioning*, edge weight = 0.067, IQR = [0.055, 0.081]; *intrinsic motivation for exercise*, edge weight = 0.037, IQR = [0.025, 0.050]; *environment*, edge weight = 0.072, IQR = [0.057, 0.083]); and *mHealth technology use*, edge weight = 0.092, IQR = [0.078, 0.104]. Two nodes showed negative, direct associations with PA: *helping relationships*, edge weight = −0.049, IQR = [−0.061, −0.035]; and *external regulation for exercise*, edge weight = −0.058, IQR = [−0.070, −0.043]. Panel C represents the indirect associations, namely the edges related to the nodes that are directly associated with PA. This two-step-neighborhood network connects 40 nodes out of the 45 nodes in the network.

**Figure 1:**
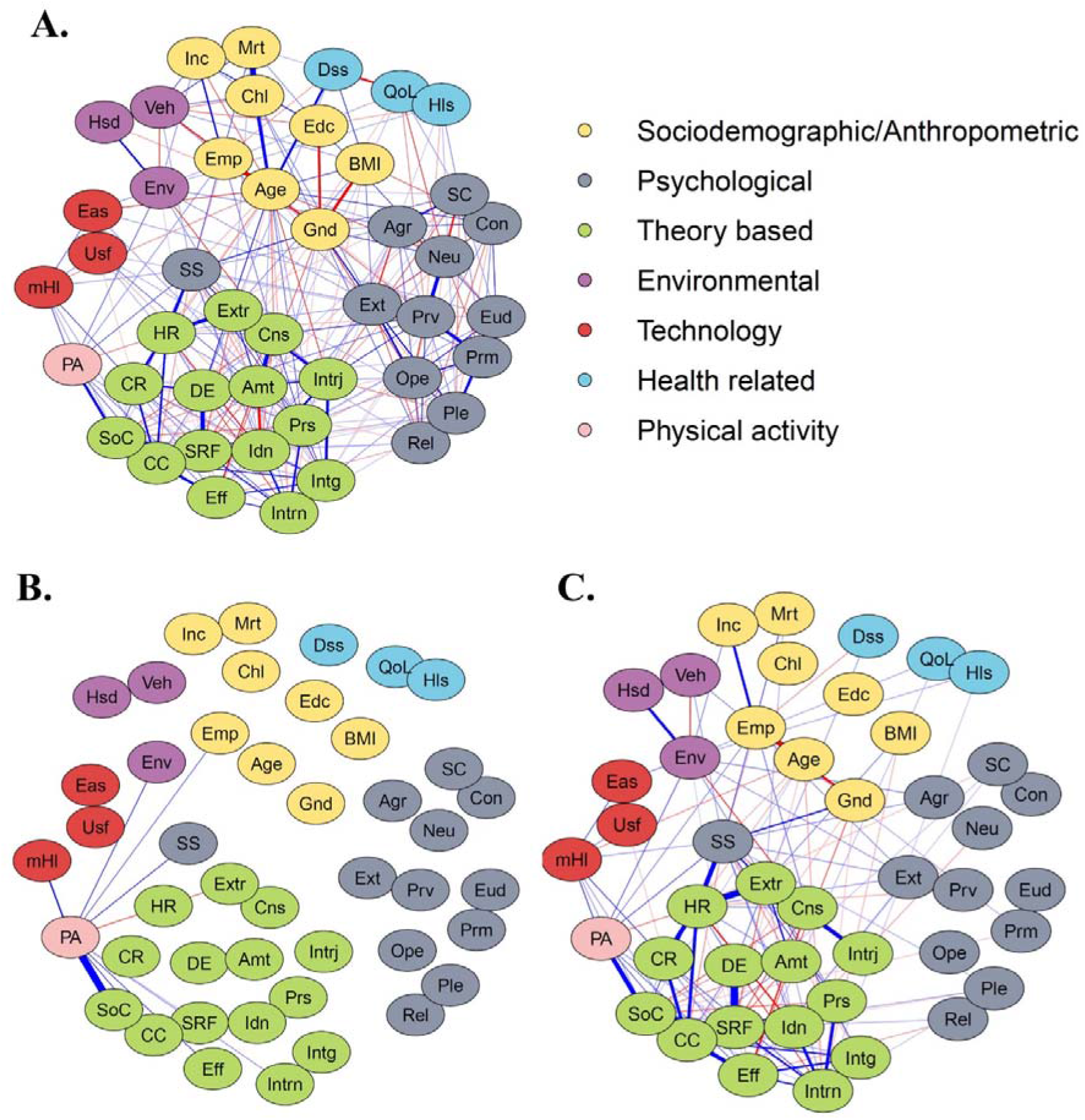
The Estimated Network of Physical Activity and its Correlates. A: The estimated network with all edges selected by the GLASSO. The thickness of each edge represents the strength of the association (partial correlation), and the colour represents the direction (blue = positive; red = negative). The node colours indicate the category of factors, corresponding to the headers in Tables 1–3. B: The network with one-step neighbourhood (displaying the edges directly associated with physical activity). C: The network with two-step neighbourhood (indirect edges added). Abbreviations and descriptions of each node are presented in Tables 1–3.

Network-centrality analyses (Figure 2) detected the highest strength for age, identified regulation, and self-reevaluation, implying that these variables had the greatest absolute associations (aggregated) with the other nodes in the network. The highest expected influence (which considers the direction of edges) was identified for integrated regulation, self-reevaluation, and promotion focus regulation, showing the greatest positive associations with the other nodes in the sum. Neuroticism exhibited the most prominent negative expected influence, which suggests that this personality dimension is typically negatively associated with the variables in the network.

**Figure 2:**
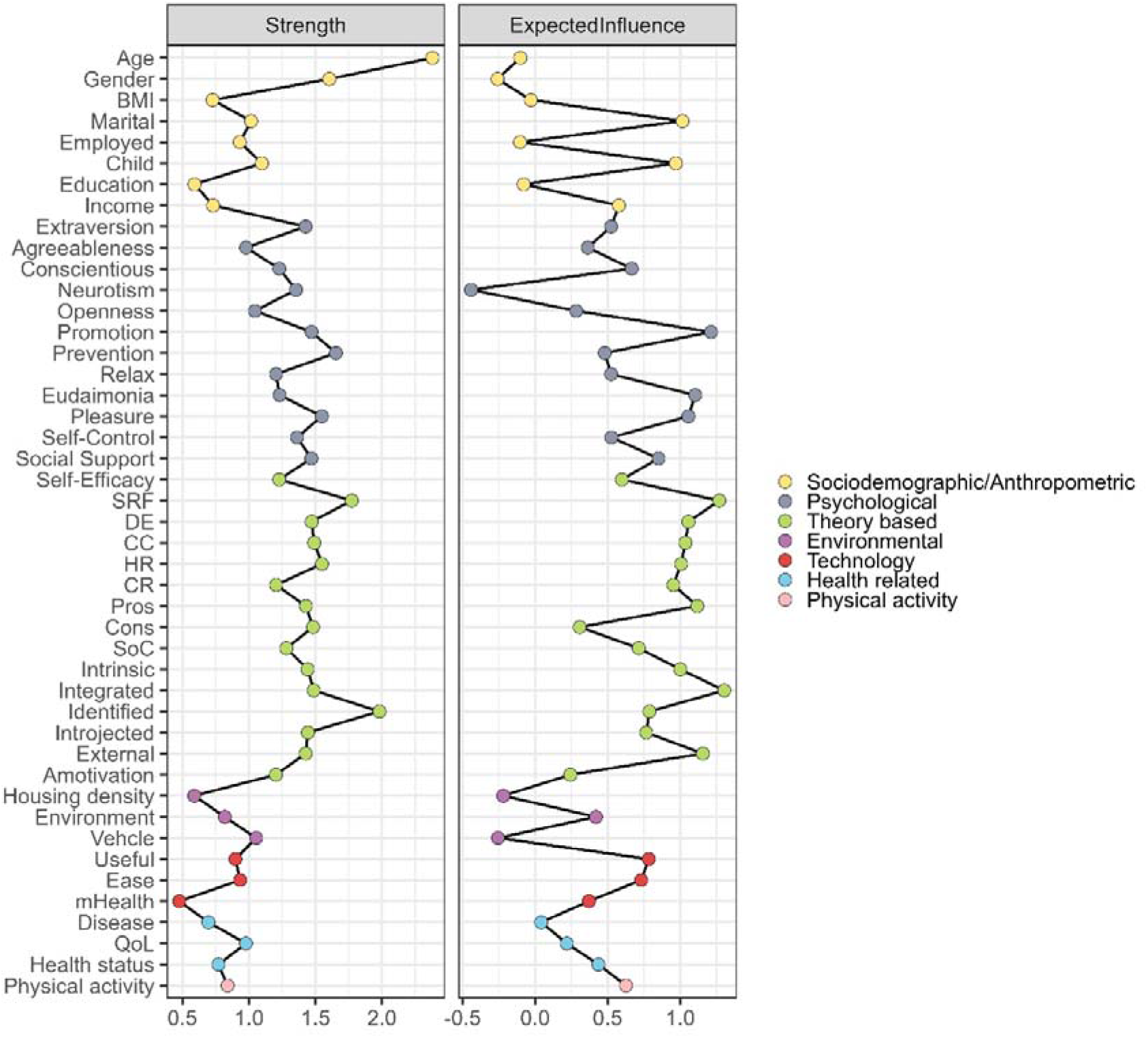
Centrality Indices. Abbreviations and descriptions of each variable are presented in Tables 1–3.

## Discussion

Insufficient physical activity has long been an unresolved health issue, and decades of research has identified numerous factors related to PA at different levels ^1,11,12,26,27^. Our aim was to reveal the factors uniquely associated with PA as well as clarify and visualise how the factors are mutually related using the network analysis. The results identified nine factors, across individual, social, and environmental levels, that are directly positively associated with PA: job/employment status, self-efficacy, perceived social support, intrinsic motivation, stage of change, counter conditioning, self-reevaluation, environment, and mHealth technology use. On the other hand, direct negative associations were identified for external regulation and helping relationships. As the edges reflect unique associations with PA after controlling for the other variables in the network, these nodes could be interpreted as proximate factors that are closely associated with PA.

The identified negative associations were intuitively difficult to interpret as both external regulation (e.g., *people around me [family, friends, doctors, etc.] say I should take up exercise*) and helping relationships (e.g., *your friends encourage you to exercise*) conceptually overlap with perceived social support (e.g., *do you have a significant other, such as a family member, spouse, friend, or colleague, who gives you advice or guidance on how to exercise?*). External regulation and helping relationships almost exclusively relate to exercise recommendations from others whereas social support is operationalised as various strategy supports that one receives, such as how-to tips, co-exercising, and appraisals. Consistent with our findings, a systematic review concluded that social pressure, like external regulation, tends to have little or even adverse effect on PA ^28^. This may suggest that the type and quality of support is significant to determine the direction of the association with PA.

Another key finding is that the two-step-neighborhood network connected almost all the variables submitted to the network analyses (40 out of 45 nodes). Here, we included as many variables as possible, following the lists of empirically and theoretically relevant correlates of PA (e.g., Trost et al. ^12^). This implies that most of the known PA-correlates are at least indirectly (if not directly) associated with PA, which reassures us that each characteristic from individual to environmental levels (e.g., demographics ^4^, motivation ^5^, self-efficacy ^6^, attitudes ^7^, and personality ^8^, environment^29^, and technology acceptance ^30^) is significant to draw a profile of someone with an (in)active lifestyle in the context of the multilevel framework ^1^.

Furthermore, the centrality analyses showed the highest strength for age, which was most closely associated with other variables in the network (followed by identified regulation and self-reevaluation). Previous studies have already identified similar associations between age and PA-correlates: e.g., personality ^31^, exercise motivation ^32^, barriers ^33,34^, and mobile device use ^35^. Conversely, environmental barriers have been documented commonly across different age groups ^34^. Interestingly, age did not have a direct edge to PA in the estimated network – individual and social characteristics may differ across age groups, but age per se works as a pure mediator, not directly informing PA levels after controlling for other PA-correlates. The highest expected influence was found for integrated regulation, followed by self-reevaluation and promotion focus regulation, which had mostly positive associations with other nodes in the network. On the other hand, neuroticism showed the greatest negative expected influence. These findings imply that motivation and process of change may play—as the transtheoretical model and self-determination theory assumed—key roles in active lifestyles whereas neuroticism and mental health issues could be a barrier in maintaining individual and environmental factors supporting PA—indeed, neuroticism is related to exercise barriers ^36^.

Several significant limitations should be noted when interpreting our results. First, the cross-sectional nature of the study limits the ability for causal inference. Bauman et al. ^1^ emphasised the significance of identifying determinants, beyond correlates, to develop and establish an intervention that helps efficiently increase engagements in PA. Although we believe that our findings refined the list of PA-related factors by identifying the direct (and indirect) associations, a longitudinal study is warranted to explore potential prospective (and causal) associations among the correlates. Second, we aimed to analyse PA-related variables as comprehensively as possible, but exhaustive assessments with *any* relevant measures were not possible for pragmatic reasons. Critically, the network structure may vary depending on the variables submitted to the analysis, which calls for careful replication with different selections of variables. Third, we solely relied on self-report measures, which may be affected by reporting bias. It would be significant to apply objective assessment methods (e.g., accelerometers) in future replications.

## Conclusion

Notwithstanding these limitations, we believe that our findings make meaningful contributions to the literature – (a) we identified 11 factors directly associated with PA among the 44 factors identified relevant in previous studies; and (b) we visualised the complex mediational associations between the PA correlates. Most of the PA correlates—from individual to environmental—were indirectly associated with PA, which confirms the significance of the multilevel perspective in understanding the contexts that facilitate active lifestyles. Another unique contribution is that our sample included only Japanese-speaking adults. This may limit the generalisability of our findings to Western populations but may support the validity of the theories and empirical findings (e.g., the transtheoretical model) in the East. It would be an interesting direction for future research to identify a unity theory for PA correlates and determinants as well as explore culture- or region-specific factors contributing to PA.

## Authors contributions

All authors contributed to the study design. **Takeyuki Oba**: Methodology, Data curation, Formal Analysis, Writing – Original draft preparation. **Keisuke Takano**: Methodology, Data curation, Supervision, Writing – Reviewing and Editing. **Kentaro Katahira**: Writing – Reviewing and Editing. **Kenta Kimura**: Project administration, Writing – Reviewing and Editing

## Data sharing

Data cannot be shared publicly because consent for data sharing was not obtained from each participant. Data is available from the corresponding author for researchers who meet the criteria for access to confidential data, which is granted by the ethics committee of the National Institute of Advanced Industrial Science and Technology.

## Patient and public involvement

Patients and/or the public were not involved in the design, or conduct, or reporting, or dissemination plans of this research.

## Ethical approval

This study was approved by the Ethics Committee of the National Institute of Advanced Industrial Science and Technology (Approval ID:2022-1279).

