## Supplemental file for "Exploring Individual, Social, and Environmental Factors Related to Physical Activity: A Network Analysis"

**Supplemental Information**

**1. Measures**

***International Physical Activity Questionnaire Short Form***

The International Physical Activity Questionnaire Short Form ^1,2^ (IPAQ-SF) was used to assess physical activity (PA) levels (see Lee et al. ^3^ for the validation against objective measures). The IPAQ-SF considers the following three activity categories: (a) walking, (b) moderate-intensity activity, and (c) vigorous-intensity activity (sedentary behaviour was not used for the current analyses). For each category, participants reported the number of days and minutes per day devoted to the activity over an average week. The weekly minutes devoted to each category were calculated by multiplying the reported number of days and minutes per day, which were then transformed into metabolic equivalents (METs) to represent total PA.

***Stage-of-Change Questionnaire***

Readiness for PA was assessed using the Japanese version ^4,5^ of the stage-of-change questionnaire ^6,7^. Participants indicated the most applicable statement among the following five: *I currently do not exercise and do not intend to start exercising in the future* (precontemplation*); I currently do not exercise but I am thinking about starting to exercise in the next six months* (contemplation); *I currently exercise some, but not regularly* (preparation); *I currently exercise regularly, but have only begun doing so within the last six months* (action); *I currently exercise regularly and have done so for longer than six months* (maintenance). Regular exercise was explicitly defined as exercising twice or more times per week for 20 minutes or longer. In the network analysis, stages were coded as precontemplation = 1 to maintenance = 5.

***The 5-level EQ-5D Version***

Each participant indicated their health state by selecting the most appropriate statement (*no problems – extreme problems*) in each of the five dimensions: mobility, self-care, usual activities, pain/discomfort, and anxiety/depression. These responses resulted in a 5-digit code that described the participant’s health state, which was converted into a numeric variable, namely the Quality of Life (QoL) score ^8^. Following this QoL part, participants were asked to indicate their subjective health today, using a visual analog scale (slider) ranging from 0 (the worst condition that the participant can imagine) to 100 (the best health condition that they can imagine).

***Ten-Item Personality Inventory***

The Big-five personality traits ^9^ were assessed using the Japanese version of the 10-item personality inventory (TIPI-J) ^10,11^. Each personality dimension comprises two adjectival items from among the following: openness to experience (*open to new experience* and *complex*), conscientiousness (*dependable* and *self-disciplined)*, extraversion (*extraverted* and *enthusiastic*), agreeableness (*sympathetic* and *warm*), and neuroticism (*anxious* and *easily upset*). Participants indicated how applicable each item was to themselves, using a 7-point scale (*Not at all* = 1; *Very much* = 7). The mean score of two items was calculated for each personality dimension.

***Regulatory Focus Questionnaire***

Two types of regulatory focuses, promotion and prevention focuses, were assessed using the Japanese version of the regulatory focus questionnaire ^12,13^. These two focuses are characterised by approach and avoidance motivation, respectively. We omitted two items related to school attitudes, as our study targeted adults. Consequently, both the promotion focus subscale (e.g., *I frequently imagine how I will achieve my hopes and aspirations*) and prevention focus subscale comprised seven items (e.g., *In general, I am focused on preventing negative events in my life*). Participants indicated how applicable each item was to themselves, using a 7-point scale (*Not at all true of me* = 1; *Very true of me* = 7). Total scores were calculated for each subscale. Cronbach’s alphas in the current data were 0.86 for the promotion focus subscale and 0.81 for the prevention focus subscale, respectively.

***Hedonic and Eudaimonic Motives for Activity Scale***

The Japanese version of hedonic and eudaimonic motives for activity ^14,15^ (HEMA) scale was used to assess motivations that contribute to participants’ sense of well-being. In the Japanese version, the hedonia subscale of the original version was further divided into two separate subscales based on arousal level: pleasure (high arousal) and relaxation (low arousal). Thus, the scale comprises three subscales: eudaimonia (e.g., *Seeking to use the best in yourself*), pleasure (e.g., *Seeking fun*), and relaxation (e.g., *Seeking to take it easy*). Participants indicated how applicable each item was to themselves, using a 7-point scale (*Not at all* = 1; *Very much* = 7). Total scores were calculated for each of the three subscales. Cronbach’s alphas in the current data ranged from 0.86 to 0.88.

***Brief Self-Control Measure***

Self-control—the ability to execute goal-oriented behaviour despite the presence of temptations to perform otherwise—is assessed using the Japanese version of the brief self-control measure (BSCM)^16,17^. This scale comprises 13 items (e.g., *I am good at resisting temptation*), including nine reverse-scored items. Participants indicated how applicable each item was to themselves, using a 5-point scale (*Not at all* = 1; *Very much* = 5). Responses to reverse-scored items were flipped and a total score was calculated. This scale had good reliability for the current data (Cronbach’s alpha = 0.85).

***Social Support Scale***

We applied the social support scale developed by Oka et al. ^18^ based on the work of Eyler et al.^19^ and Sallis et al.^20^. This scale includes five items, each relating to whether significant others in participants’ lives, such as family, spouses, friends, and colleagues, fit the following descriptions: (a) *gave me advice and guidance on how to exercise*, (b) *gave me time to exercise*, (c) *gave me encouragement or reinforcement for exercising*, (d) *exercise with me*, and (e) *gave me rewards for exercising*. Participants responded with ‘yes’ or ‘no’ for each item. The total score was calculated with ‘yes’ responses scored as 1 and ‘no’ as 0. Cronbach’s alpha was 0.74 for the current data.

***Self-Efficacy Scale***

The Japanese version of the Self-Efficacy Scale was administered ^4,21^. Participants indicated their confidence level in performing PA and exercise in the presence of specific barriers: (a) when being physically exhausted, (b) not motivated for PA, (c) busy with other things, and (d) in bad weather. These four items were rated on a 5-point scale (*Not at all* = 1; *Very much* = 5). The cumulative total score showed excellent reliability (Cronbach’s alpha = 0.91) for the current data.

***Decisional Balance Scale***

The perceived positive and negative aspects of PA were examined using an adapted version ^22^ of the original scale ^23^ with items from other scales related to decisional balance ^24,25^. This scale has 20 items and divides into the two subscales: *pros* and *cons*. Each subscale comprises 10 items (e.g., *I would have more energy for my family and friends if I exercised regularly*). Participants rated how well each item described their way of thinking and attitudes toward PA and exercise on a 5-point scale (*Not at all* = 1; *Very much* = 5). One of the *pros* items (*Regular exercise helps lose weight and gain strength*) was erroneously excluded from the current survey. Therefore, we calculated the mean scores of nine items for *pros* and 10 items for *cons* (Cronbach’s alpha = 0.86 for *pros*; 0.86 for *cons*).

***Process-of-Change Questionnaire***

We used the 21-item version of the process-of-change questionnaire ^26^. This scale has the following five factors: (a) self-revaluation, reinforcement management, and self-liberation (seven items; e.g., *You feel more confident when you exercised regularly*); (b) dramatic relief and environmental reevaluation (five items; e.g., *You get upset when you saw people who would benefit from exercise but chose not to exercise*); counter conditioning (three items; e.g., *You feel tired, and made yourself exercise anyway because you knew you would feel better afterwards*); helping relationships (three items; e.g., *You have someone around who encouraged you to exercise*); and consciousness raising (three items; e.g., *You read articles to learn more about exercise*). Each item was rated on a 5-point scale (*Never* = 1; *Repeatedly* = 5) and the cumulative total score was calculated for each process-of-change factor. Cronbach’s alpha ranged from 0.72 to 0.91 for the current data.

***Revised Self-determined Motivation Scale for Exercise***

Exercise motivation was assessed using the revised self-determined motivation scale for exercise (SMSE-2) ^27^. The SMSE-2 has 22 items with six subscales representing the key constructs of the self-determination theory ^28^. The subscales include intrinsic motivation (four items; e.g., *Exercising itself is fun*), integrated regulation (four items; e.g., *Doing exercise and being myself are inseparable*), identified regulation (four items; e.g., *It is important to me to exercise*), introjected regulation (four items; e.g., *I feel guilty if I do not exercise*), external regulation (three items; e.g., *I exercise because other people will be pleased with me*), and amotivation (three items; e.g., *I do not know why I exercise*). Each item was rated on a 5-point scale (*Not at all true* = 1; *Very true* = 5). The cumulative total scores for each subscale showed good internal consistencies (Cronbach’s alpha = 0.78-0.88).

***International Physical Activity Questionnaire—Environmental Module***

The International Physical Activity Questionnaire—Environmental Module ^29^ (IPAQ-E) was used to assess physical environmental factors for PA participation. Following the validation study among a Japanese sample ^29^, 17 items were administered, which were scored for three domains: (1) residential density, (2) environmental conditions, and (3) the number of household motor vehicles. The residential density was indicated by selecting the most applicable housing type for participants’ neighbourhood (e.g., *detached single-family residences*; *townhouses, row houses, apartments, or condos of 2-3 stories*). The environmental conditions comprised 15 items, encompassing presence of sidewalks, bike paths, and recreational facilities as well as aesthetic, social, and safety attributes ^30^. Each item was rated on a 4-point scale (*Strongly disagree* = 1; *Strongly agree* = 4). The cumulative score was calculated. Regarding household motor vehicles, participants responded to: how many motor vehicles are there in your household?

***Use of mHealth Technology***

We asked participants whether they were using any smartphone applications or activity trackers supporting PA and exercise. Participants gave a yes–no binary response, and upon an affirmative response, they provided further details about how they were using the mobile devices (e.g., duration, frequency). For the current network analysis, we used only the binary response. Other detailed responses were analysed and reported elsewhere ^31^.

***Technology Acceptance***

Perceived usefulness (e.g., *work more quickly*) and perceived ease of use (e.g., *easy to learn*), were assessed as key dimensions of technology acceptance ^32,33^. The Japanese version comprises six items (three items each for perceived usefulness and ease of use), rated on a 7-point scale (*Strongly disagree* = 1; *Strongly agree* = 7). The cumulative total scores were calculated. Cronbach’s alphas were 0.89 for perceived usefulness and 0.88 for perceived ease of use.

***Present Disease***

Participants indicated whether they had any disease at the time of the survey (yes–no binary response). Upon an affirmative response, they further indicated any applicable disease/s on a list of (mostly non-communicable) disease names. Table S1 shows the frequency of each disease reported in the present sample. Hypertension was the most prevalent, followed by hyperlipidemia and diabetes. The presence (vs. absence) of any disease, instead of the presence of each specific disease, was submitted to the network analysis. This decision was made because most of the diseases were not prevalent (< 5%) and we wanted avoid data imbalance for network estimation.

Table S1: Reported Diseases and Frequency

| Disease | *N* (%) |
| --- | --- |
| Asthma | 518 (2.73%) |
| Stroke (excluding cerebral hemorrhage / infarction) | 33 (0.17%) |
| Cerebral hemorrhage | 59 (0.31%) |
| Cerebral infarction | 116 (0.61%) |
| Other cerebrovascular diseases | 66 (0.35%) |
| Chronic renal failure | 84 (0.44%) |
| Dialysis | 50 (0.26%) |
| Angina pectoris | 204 (1.07%) |
| Myocardial infarction | 103 (0.54%) |
| Heart failure | 72 (0.38%) |
| Ischemic heart disease | 26 (0.14%) |
| Other cardiac diseases | 127 (0.67%) |
| Anemia | 424 (2.23%) |
| Hypertension | 3390 (17.84%) |
| Low blood pressure | 130 (0.68%) |
| Arrhythmia | 397 (2.09%) |
| Hyperlipidemia | 1745 (9.18%) |
| Hyperuricemia | 374 (1.97%) |
| Renal diseases other than renal failure | 102 (0.54%) |
| Diabetes mellitus | 1078 (5.67%) |
| Liver and kidney disease | 52 (0.27%) |
| Gastric and duodenal ulcer | 115 (0.61%) |
| Depression | 573 (3.01%) |
| Osteoporosis | 585 (3.08%) |

**2. Sensitivity Analysis**

Isvoranu and Epskamp ^34^ suggested using Spearman correlations as an input for network estimation in the case of data skewness (as an alternative approach for paranormal transformation). Following this recommendation, we estimated a partial correlation network using Spearman correlations. The results were almost identical as the network estimated with a non-paranormal transformation (reported in the main manuscript), except for age as an additional node directly associated with PA (Figure S1). The estimated edge weights (direct associations with PA) were as follows: *age*, edge weight = 0.038, IQR = [0, 0.052]; *job/employment*, edge weight = 0.052, IQR = [0.038, 0.064]; *stage of change*, edge weight = 0.276, IQR = [0.262, 0.290]; *social support*, edge weight = 0.074, IQR = [0.059, 0.088]; *self-efficacy*, edge weight = 0.054, IQR = [0.041, 0.067]; *self-reevaluation*, edge weight = 0.028, IQR = [0, 0.043]; *counter conditioning*, edge weight = 0.069, IQR = [0.056, 0.082]; *helping relationships*, edge weight = -0.050, IQR = [-0.062, -0.036]; *intrinsic motivation for exercise*, edge weight = 0.042, IQR = [0.030, 0.054]; *external regulation for exercise*, edge weight = -0.055, IQR = [-0.067, -0.041]; *environment*, edge weight = 0.075, IQR = [0.061, 0.088]; and *mHealth technology use*, edge weight = 0.096, IQR = [0.082, 0.109].

Figure S1: The Network Estimated Based on Spearman Correlations

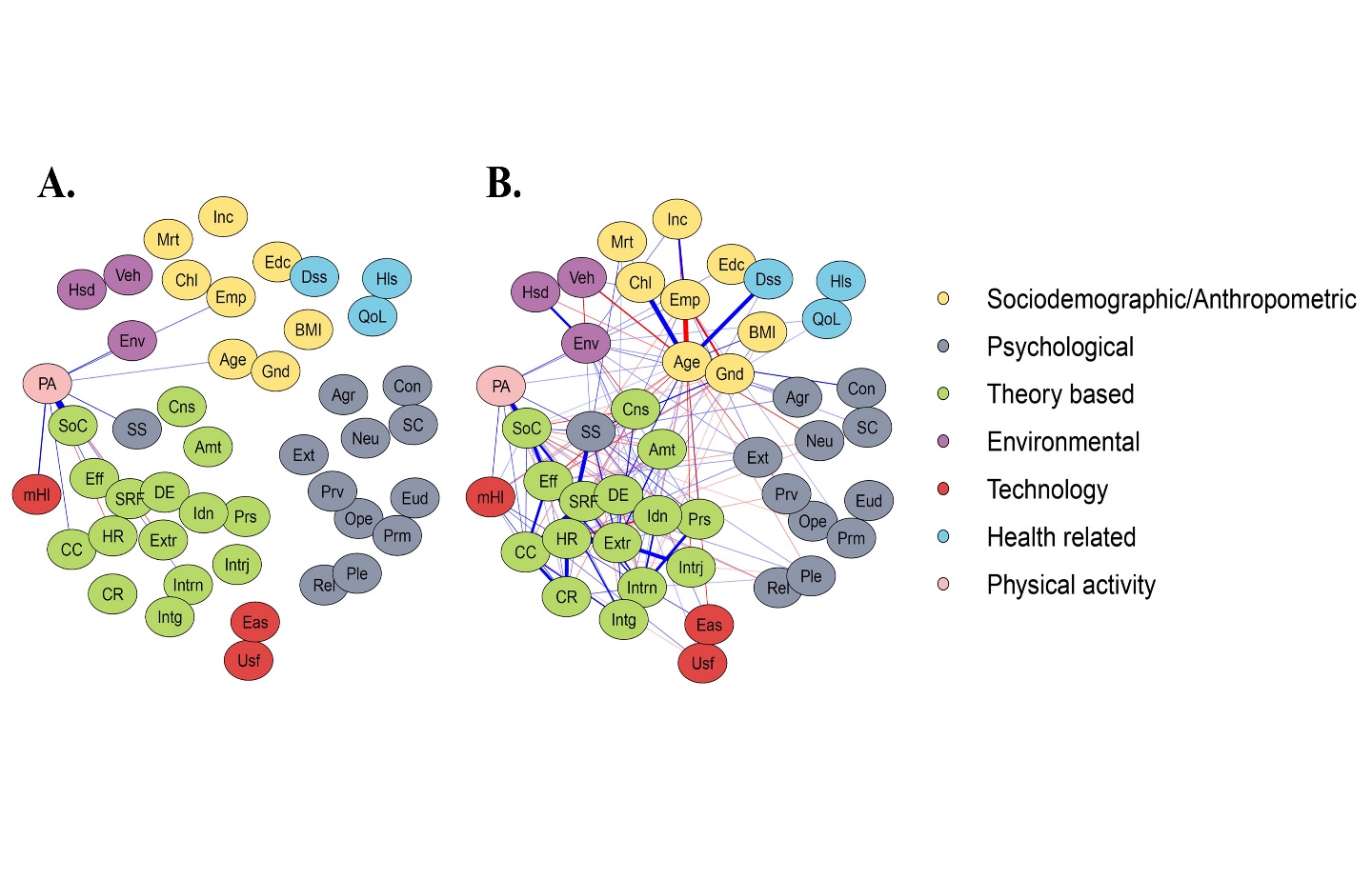

A: The estimated network with one-step neighbourhood (displaying the edges directly associated with PA). B: The network with two-step neighbourhood (indirect edges added). Abbreviations and descriptions of each node are presented in Tables 1–3 in the main text.

**3. Accuracy**

Network accuracy was assessed with confidence intervals (CIs) for each edge weight. Bootstrap resampling (2,500 iterations) was conducted using the *bootnet* package ^35^ in R. Following Epskamp et al. ^35^, we computed 2.5% and 97.5% quantiles of each edge estimate, which are interpreted as the approximate upper and lower limits of a 95% CI. Figure S2 shows the point estimates and CIs for each edge selected by the GLASSO. Edges with CIs including zero should be interpreted with caution as these edge estimates might not be sufficiently robust (or might not be selected in a stable manner). Of the 295 GLASSO-selected edges, 220 edges (74.6%) did not include 0 in the CIs, implying that most of the edges (and thus, the overall network) could be considered accurate.

Figure S2: Edge Accuracy (Edge Weight Estimates and Confidence Intervals)

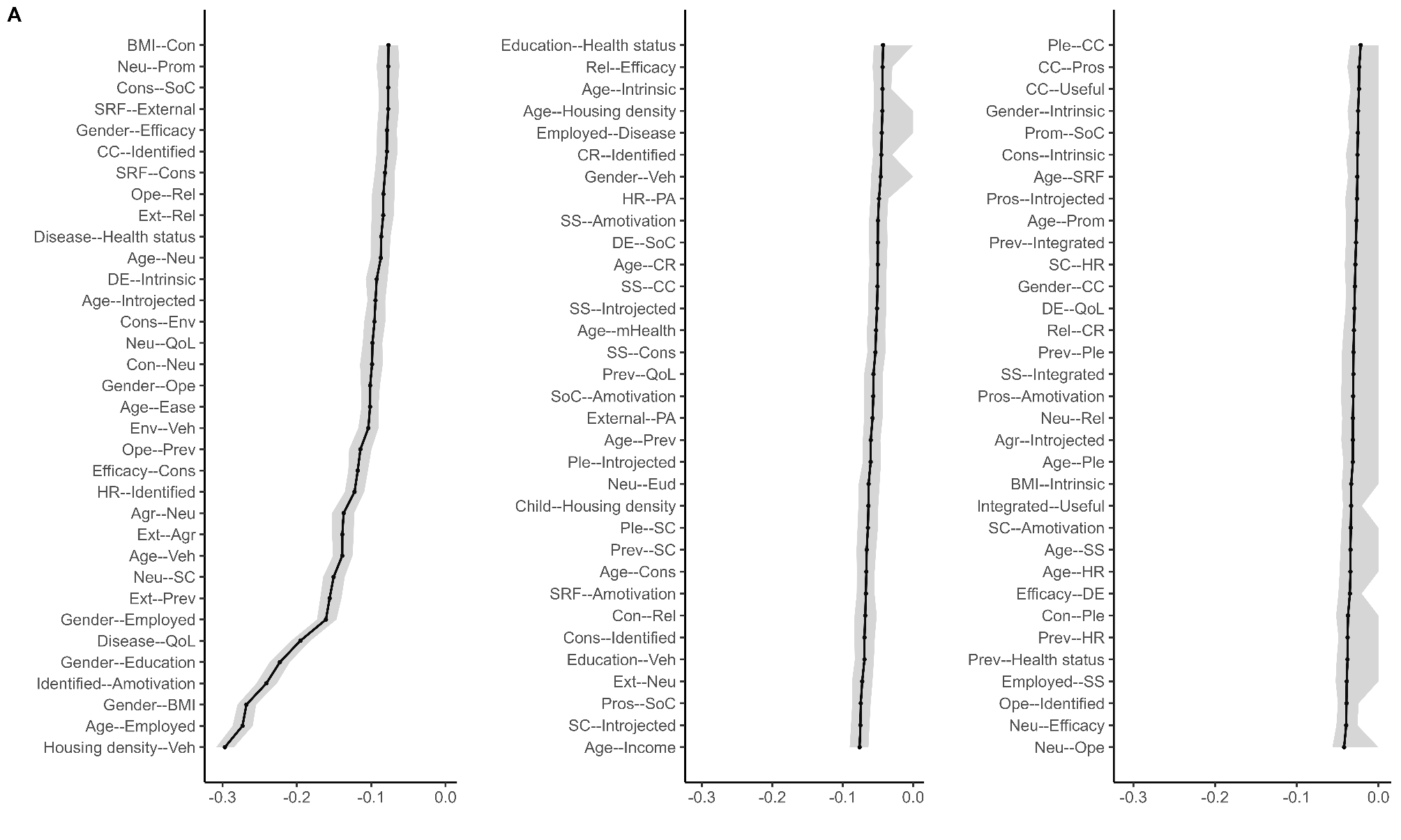

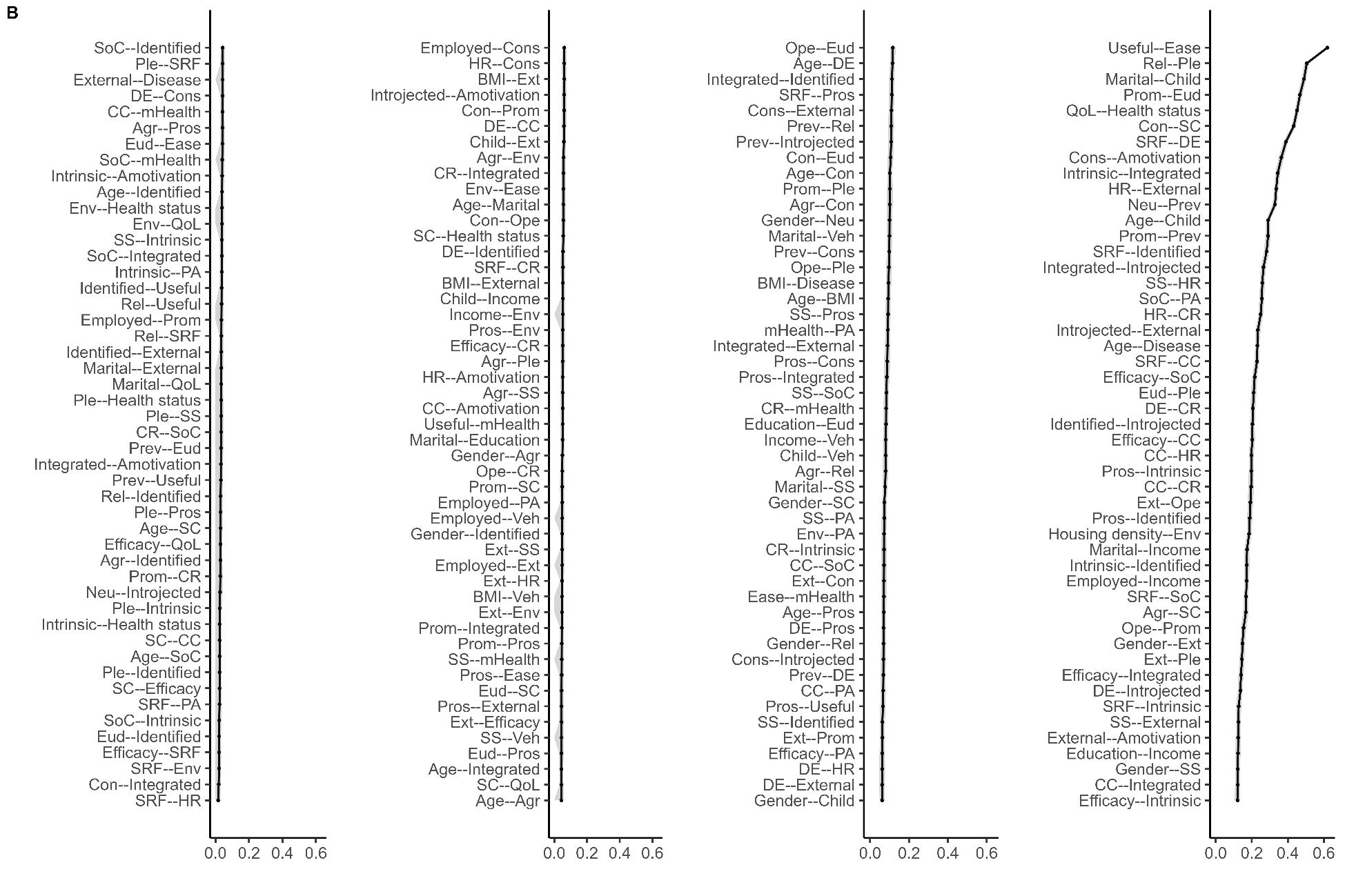

All GLASSO-selected edges are displayed (A: Negative edges; B: Positive edges). Shaded areas indicate CIs with the 2.5th and 97.5th quantiles.

**4. Stability**

The stability of each centrality index was assessed with correlation stability (CS) coefficients. A CS coefficient represents how many cases can be dropped from the data to retain the centrality in the original network (with a cutoff of > 75%) ^35^ while maintaining a correlation (*r* > 0.7) between the centrality values estimated based on the original data and those computed with less cases. Additionally, a CS coefficient of 0.5 is the minimum to interpret differences in centrality between nodes ^35^. Figure S3 shows CS coefficients (for strength and expected influence) as a function of the amount of data sampled from the original data. Even 30% of sampling maintained excellent CS coefficients, greater than 0.95.

Figure S3

*Centrality Stability*

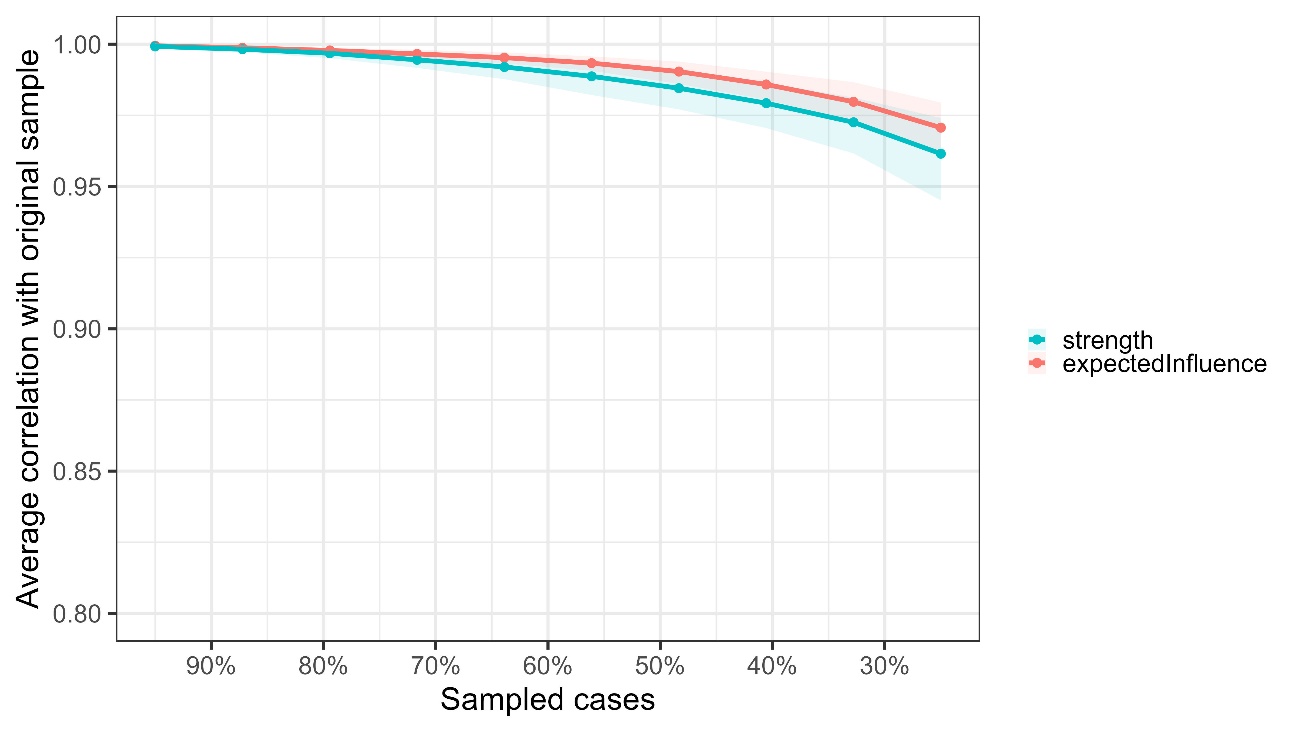
